# Serological Cytokine and chemokine profile in pregnant women with COVID19 in Mexico City

**DOI:** 10.1101/2020.07.14.20153585

**Authors:** A. Cérbulo-Vázquez, B. Zavala-Barrios, JC Briones-Garduño, GML Guerrero-Avendaño, L. Arriaga-Pizano, E. Ferat-Osorio, L., G.L. Cabrera-Rivera, P. Miranda-Cruz, M.T. García de la Rosa, J.L. Prieto-Chavez, V. Rivero-Arredondo, R.L. Madera-Sandoval, A. Cruz-Cruz, E. Salazar-Rios, D. Serrano-Molina, R. C. De Lira-Barraza, A. H. Villanueva-Compean, A. Esquivel-Pineda, R. Ramirez-Montes de Oca, G. Flores-Padilla, C. López-Macías

## Abstract

On January 30th, 2020, the WHO declared the outbreak of COVID19, a disease due to the new coronavirus called SARS-CoV-2. Certain comorbidities, symptoms and signs are characteristic of COVID19 in the general population and in pregnant women. However, pregnant women are considered as a high-risk group for COVID19. To know about the frequency of comorbidities, signs and symptoms, the presence of lymphopenia, antibodies response to SARS-CoV2 and cytokine and chemokine serum concentration, six pregnant women with COVID19 were studied at the moment of admission. The lower concentration of CCL17 was detected in the Pregnant COVID19 group, similar concentration of IL-6 was also detected in non-pregnant and pregnant COVID19 patients. Our result show that pregnant and non-pregnant women with COVID19 has similar cytokine profile.

## Introduction

On January 30th, 2020, the WHO declared the outbreak of COVID19 disease due to a new coronavirus called SARS-CoV-2 (1). Certain comorbidities are important to consider for severity in COVID19. Hypertension, cardiovascular disease, diabetes and obesity are comorbidities that increase the risk for mortality and severity (2, 3). Some symptoms and signs are frequently recognized in COVID19 patients, among them fever, cough and dyspnea are common (2, 4). Also, an extensive cellular infiltration of neutrophils and macrophages into the pulmonary interstitium and alveoli is observed in COVID-19 patients (5, 6). Furthermore, an increased number of neutrophils and monocytes in blood is detected, in contrast a lymphopenia has been observed in patients with a fatal SARS and COVID19 outcome (7-10), in addition, an extensive SARS-CoV-1-induced apoptosis has been detected in respiratory epithelial cells, neurons, T lymphocytes and dendritic cells (11, 12).

Humoral response against SARS-CoV-2 is detected as early as three or four days after symptoms onset (13-15). IgM seroconversion rate reach its peak around 20-22 days after symptoms onset, and IgG peak is around 17 to 19 days after symptoms onset (13). The real-time reverse transcription-polymerase chain reaction (RT-PCR) of nasopharyngeal swabs for SARS-Co-2 is the gold standard for diagnosis (16). Also, some other parameters such as the Neutrophil count would be useful to COVID19 diagnosis (17).

The infection with CoV upregulate the expression of cytokines and chemokines. Infection of Dendritic Cells (DC) with SARS-CoV induces low expression of IFN-α and β, and also a moderate expression of TNF-α and IL-6 with high expression of CCL3, CCL5, CCL-2 and CXCL10 (18). Serum high concentrations of proinflammatory cytokines and chemokines with low IL-10 concentrations have been detected in patients with SARS-CoV (19). In addition, it has been reported a CCL2 and CXCL8 increase synthesis *in vitro* by protein S of the SARS-CoV (20, 21). Regarding to COVID19, cytokine and chemokine response are also detected in SARS-CoV2 infected patients, and treated in Intensive Care Unit (ICU). In non-ICU patients has been detected a higher concentration of IL-2, IL-7, IL-10, G-CSF, IP10 (CXCL-10), MCP-1 (CCL2), MIP1a (CCL3) and TNF-α than in control patients (4). SARS-CoV-2-induced cytokines and chemokines could have several sources besides lung epithelial cells. CCL2 is produced by many cell types, including endothelial, and epithelial, but the monocytes/macrophages are the major source (22), macrophages are also the primary source of IL-1 (23) and CXCL10 in response to virus infection (24). IL-6 is expressed by endothelial cells in response to hypoxia (25), and the IFN-γ is major expressed by NK and NKT cells (26).

For SARS-CoV emergency in 2004, the largest series of pregnant patients had a case-fatality rate of 25% (27), and by this reason maternal death could be high in COVID19 pregnant women. The immunological condition in pregnancy is complex and involves series of mechanism that regulates the immune response. The maternal immune system expresses a tolerance state that allows the progression of pregnancy and perpetuation of specie. Despite the high virulence of COVID19 with case-fatality rate around 5 to 6 % in general population, no maternal deaths from SARS-CoV-2 infection have been reported (28, 29). An exacerbated inflammatory response in pregnancy could affect the delicate immune balance and complicate the normal progression. To know about the frequency of comorbidities, signs and symptoms, the presence of lymphopenia, antibodies response to SARS-CoV-2 and cytokine and chemokine serum concentration, pregnant women with COVID19 were compare with non-pregnant COVID19 women, and non-pregnant COVID19 negative women.

## Material and methods

### Patients

This study was jointly conducted by the Medical Research Unit on Immunochemistry (UIMIQ), Specialties Hospital, National Medical Center “ XXI Century” and Gynecology & Obstetrics Department in the General Hospital of Mexico “ Dr. Eduardo Liceaga” (Research project: R-2020-785-095). Eighteen women were enrolled after obtaining a signed informed consent. Three subgroups were analyzed as follows: a) Non-pregnant, Non-COVID19 women (Non-COVID19, n=7), Female Medical Doctors who care COVID19 patients integrates this group; b) confirmed SARS-CoV2-infected Non-Pregnant women (NP-COVID19, n=5); and c) confirmed SARS-CoV-2-infected Pregnant women (P-COVID19, n=6). SARS-CoV2 viral infection was confirmed by specific reverse transcription–polymerase chain reaction (RT-PCR) as ministry of health in México indicated (30). The diagnosis of COVID19 was based on clinical characteristics (31), comorbidities and clinical signs or symptoms were registered and shown in Table 1 and 2 respectively.

**Table 1.** Clinical characteristics of pregnant and non-pregnant women with or without COVID19.

|  | **Non-COVID19**<br>**(n=7)** | **NP-COVID19**<br>**(n=5)** | **P-COVID19**<br>**(n=6)** | **p** |
| --- | --- | --- | --- | --- |
| **Age** | 27.3±1.8 | 28.4±3.2 | 33.7±6.2 | 0.145 |
| **Weight (Kg)** | 66.5±8.5 | 72.4±23.6 | 76±11.9 | ns |
| **Height (m)** | 1.58±0.05 | 1.56±0.1 | 1.59±0.04 | ns |
| **Respiratory Rate**<br>**(breaths per min)** | 19.7±4 | 24.2±4.9 | 23.3±6.1 | ns |
| **Heart Rate**<br>**(beats per min)** | 74±9.5 | 109±15.5 | 105.3±11.1 | **0.009<sup>a</sup>**<br>**0.014<sup>b</sup>** |
| **Temperature (°C)** | 35.8±0.6 | 36.7±0.8 | 37±0.5 | **0.020** |
| **Diastolic blood**<br>**pressure (mmHg)** | 102.1±9 | 117.2±10.9 | 112.2±10.4 | ns |
| **Systolic blood**<br>**pressure (mmHg)** | 64.9±5.3 | 63.6±7.3 | 73.5±8 | ns |

Non-COVID19; Non-pregnant, non-COVID19. NP-COVID19; Non-Pregnant-COVID19. P-COVID19; Pregnant-COVID19. Kruskal-Wallis test. a; Non-COVID19 vs. NP-COVID19. b; Non-COVID19 vs. P-COVID19. Significant $p < 0.05$ .

**Table 2.**
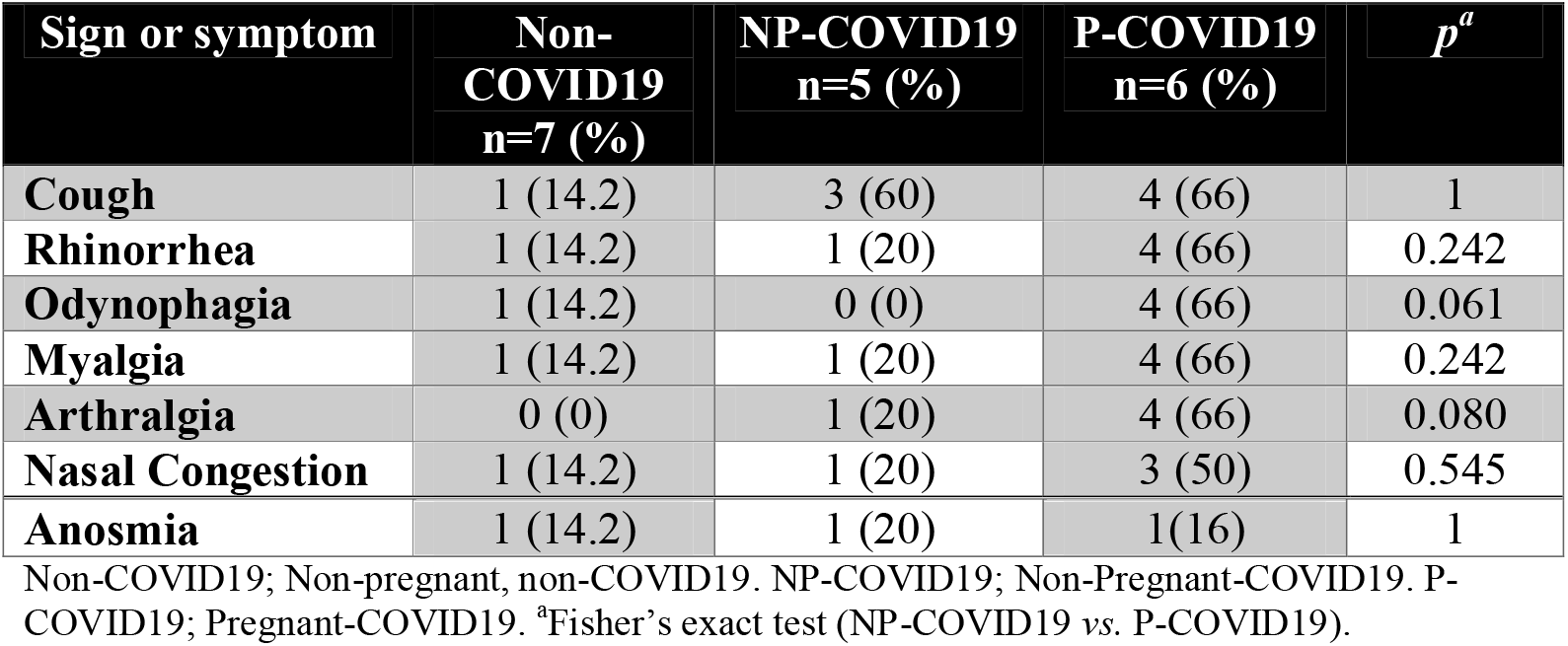
Frequency of SARS-related signs and symptoms.

| **Sign or symptom** | **Non-COVID19**<br>**n=7 (%)** | **NP-COVID19**<br>**n=5 (%)** | **P-COVID19**<br>**n=6 (%)** | **p<sup>a</sup>** |
| --- | --- | --- | --- | --- |
| **Cough** | 1 (14.2) | 3 (60) | 4 (66) | 1 |
| **Rhinorrhea** | 1 (14.2) | 1 (20) | 4 (66) | 0.242 |
| **Odynophagia** | 1 (14.2) | 0 (0) | 4 (66) | 0.061 |
| **Myalgia** | 1 (14.2) | 1 (20) | 4 (66) | 0.242 |
| **Arthralgia** | 0 (0) | 1 (20) | 4 (66) | 0.080 |
| **Nasal Congestion** | 1 (14.2) | 1 (20) | 3 (50) | 0.545 |
| **Anosmia** | 1 (14.2) | 1 (20) | 1 (16) | 1 |

Non-COVID19; Non-pregnant, non-COVID19. NP-COVID19; Non-Pregnant-COVID19. P-COVID19; Pregnant-COVID19. <sup>a</sup>Fisher's exact test (NP-COVID19 vs. P-COVID19).

### Blood Collection

Our study is in accordance with the World Medical Association’s Declaration of Helsinki. After a patient agreed to participate in the study, healthcare personal collected blood specimens in silicone-coated and heparinized tubes (BD Vacutainer, N.J, USA) samples were processed immediately after their collection. Complete blood count and serum was obtained to compares among groups.

### Determination of cytokine and chemokine levels

The serum concentrations of cytokines (IL-2, IL-4, IL-6, IL-10, TNF, IFN-γ, and IL-17) and chemokines (CXCL8/IL-8, CXCL10/IP-10, CCL11/Eotaxin, CCL17/TARC, CCL2/MCP-1, CCL5/RANTES, CCL3/MIP-1a, CXCL9/MIG, CXCL5/ENA-78, CCL20/ MIP-3a, CXCL1/GROa, CXCL11/I-TAC and CCL4/MIP-1b) were determined using bead based immunoassays (CBA kit, Cat. 560484, BD PharMingen, San Diego, CA, USA; and LEGENDplex, Cat. 740003, BioLegend, San Diego, CA, USA), according to the manufacturer’s instructions. Log-transformed data were used to construct standard curves fitted to 10 discrete points using a 4-parameter logistic model. The concentrations in the test samples were calculated using interpolations of their corresponding standard curves.

### Anti SARS-CoV-2 IgM, IgG ELISA

For IgM and IgG test, 96 well ErbaLisa kit was used (cat. IME00137 for IgM and cat. IME00136 for IgG, Erba Mannheim, London, U.K), the procedure was according to the manufacturer’s instructions. The sera from patients were inactivated at 56°C for 1 hour, and formerly incubated for 20 minutes at room temperature in the ErbaLisa plate with the SARS-Cov2 antigen. After washing the enzyme conjugate were added and incubated for 20 minutes at room temperature. Afterward, the conjugated enzyme was removed and plate were washing. The TMB substrate was added and incubated for 10 minutes at room temperature. The OD were read at 450 nm by EPOCHH (serial number 1503277, BioTek® Instruments, Inc).

Quality control was provided by the company (OD calibrator, negative control and positive control included).

### Statistical Analysis

Statistical analysis was performed using Graph Pad Prism® version 6 software (GraphPad Software, San Diego, CA, USA), and SPSS software, compilation 1.0.0.1347. Non-parametric ANOVA test (Kruskall-Wallis test) with Dunn post-test were applied. Also, categorical variables were expressed as number (%) and compared by Fisher’s exact test (NP-COVID19 vs. P-COVID19 groups). A *p*<0.05 was considered as statistically significant.

## Results

Non-pregnant non-COVID19 patients were or remaining asymptomatic for respiratory disease 14 days before and after the moment of blood sample collection. Four of five NP-COVID19 women (80%) were mild disease, and one (20%) had a critical disease and passed away, this patient looked for medical attention eleven days after symptoms onset, and required mechanical ventilation. Four P-COVID19 women delivered their newborn by cesarean, one by vaginal delivery and one had remained pregnant in her recovery COVID19 phase. At admission, the range of gestational weeks was 37 to 40 gestational weeks. Five of six P-COVID19 women (83%) were mild disease, and one (16%) had a critical disease and received non-invasive mechanical ventilation, but after treatment was discharge from service. None maternal or neonatal death was recorded in P-COVID19 group.

Table 1 shows clinical characteristics for the Non-COVID19, NP-COVID19 and P-COVID19 women. The age, weight, height, respiratory rate, diastolic, and systolic blood pressure were similar among the groups. Heart rate show a statistical difference between Non-COVID19 and NP-COVID19 or P-COVID19. Moreover, the temperature between Non-COVID19 and P-COVID19 show a statistical difference. One of the Non-COVID19 reports a Polycystic Ovary Syndrome and another hypothyroidism. One of the NP-COVID19 patients had leukemia under treatment (her hematic biometry was in normal range) and she was discharge from service after COVID19 was solved. P-COVID19 group had underlying diseases such as pre-gestational overweight/obesity (4/6), gestational diabetes (3/6), urinary tract infection (2/6), and hypothyroidism (1/6).

The most common SARS-related signs and symptoms are shown in Table 2. No statistical difference was observed for any of the data; however, the most of patients had cough, rhinorrhea, odynophagia, myalgia, and arthralgia was noted in the group of P-COVID19.

Lymphocytes, monocytes and, neutrophils absolute count were in normal range in the group of Non-COVID19. Neutropenia and neutrophilia were detected in the group of NP-COVID19, also three patients with lymphopenia were detected, and one patient had monocytosis in this group. A patient with Leukopenia in the NP-COVID19 group was recorded (3.0 ⨯ 10^3^ μL), and one patient with leukocytosis was also detected (15.6 ⨯ 10^3^ μL). For white cell count in pregnancy we take a reference range (32), and none of the P-COVID19 patients showed leukocytosis. Neutrophilia was detected in one of the P-COVID19 patients, and none lymphopenia or monocytopenia was detected. Figure 1 shown the Neutrophil/Lymphocyte Ratio, non-COVID19 group show a 2.3±0.2 mean and SD, while NP-COVID19 and P-COVID19 groups shown 16.7±18.1 and 5.9±3.9 respectively.

**Fig 1.**
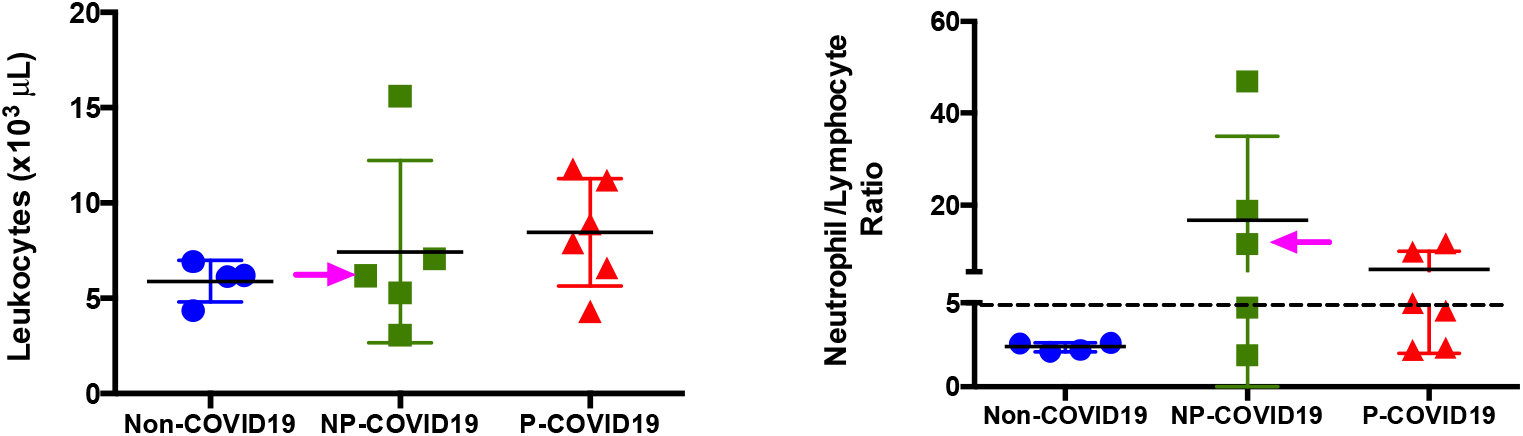
Absolut count of leukocytes and Neutrophil/Lymphocyte Ratio. Results are expressed as mean ± SD. Dash line is the cutoff for NLR. Kruskal-Wallis test. Significant *p*<0.05. The arrow points the fatal case in NP-COVID19 group.

In the group of non-COVID19 we observed one patient positive for IgM (14%) and one more for IgG test (14%). None of the NP-COVID19 patients were positive for IgM but two were for IgG (40%). Four of six of P-COVID19 patients were positive IgM test (66%), and two of six were positive for IgG (33%). A Fisher’s exact test was applied and no statistical difference was observed between IgM NP-COVID19 and IgM P-COVID19 groups (*p*=0.061), or between IgG NP-COVID19 and IgG P-COVID19 groups (*p*=1). Figure 2 show the antibody index (AI) for IgM and IgG, the AI was calculated as antibody absorbance/cutoff, the cutoff was calculated as calibrator absorbance x calibrator factor.

**Fig 2.**
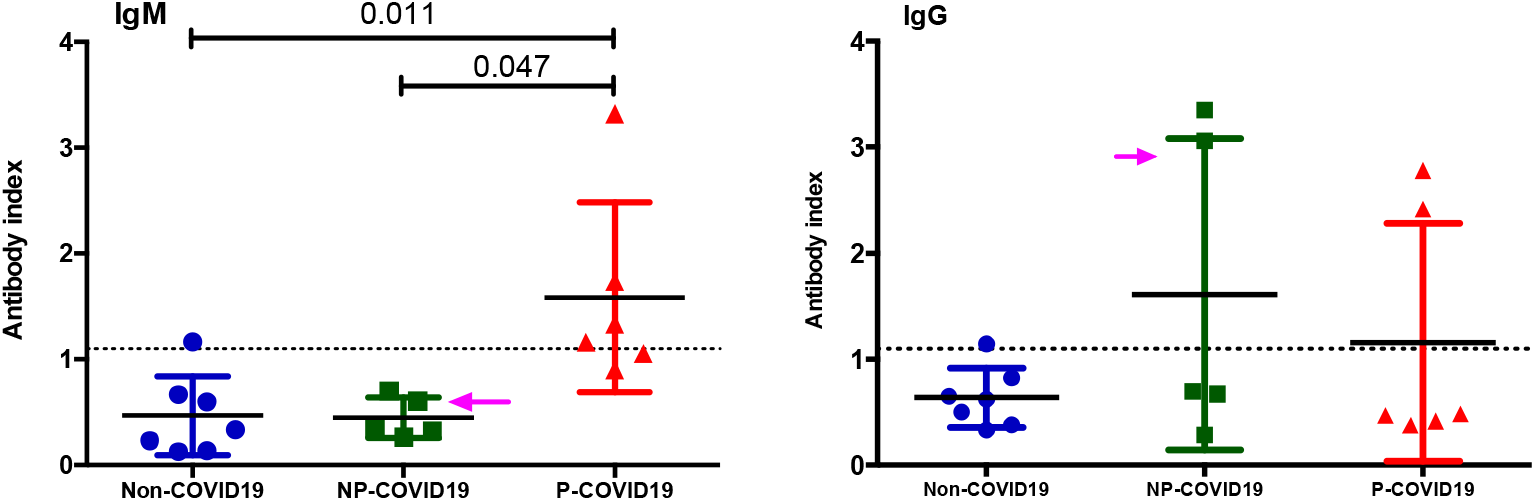
Antibody index. Results are expressed as mean ± SD. Kruskal-Wallis and Dunn’s multiple comparisons test was calculated. Significant *p*<0.05. Dot line show the cutoff value. The arrow points the fatal case in NP-COVID19 group.

Serum cytokines as IL-17, IFN-γ, CXCL8, CCL2, CCL3, CXCL10, CCL11, CXCL9, CCL20, CXCL11 and CCL4 did not show any statistically significant difference among the groups (Figure 3), only one patient in the group of P-COVID19 and one more in the NP-COVID19 expressed an IL-2 concentration above the limit of detection (9.97 and 17.45 pg/mL respectively). Either NP-COVID19 or P-COVID19 (n=11) showed TNF and IL-4 concentration below the limit of detection (<39.06 pg/mL). Figure 4 shows the cytokines and chemokines (IL-10, IL-6, CCL5, CCL17, CXCL5 and CXCL1) that showed a statistical difference among the groups.

**Fig 3.**
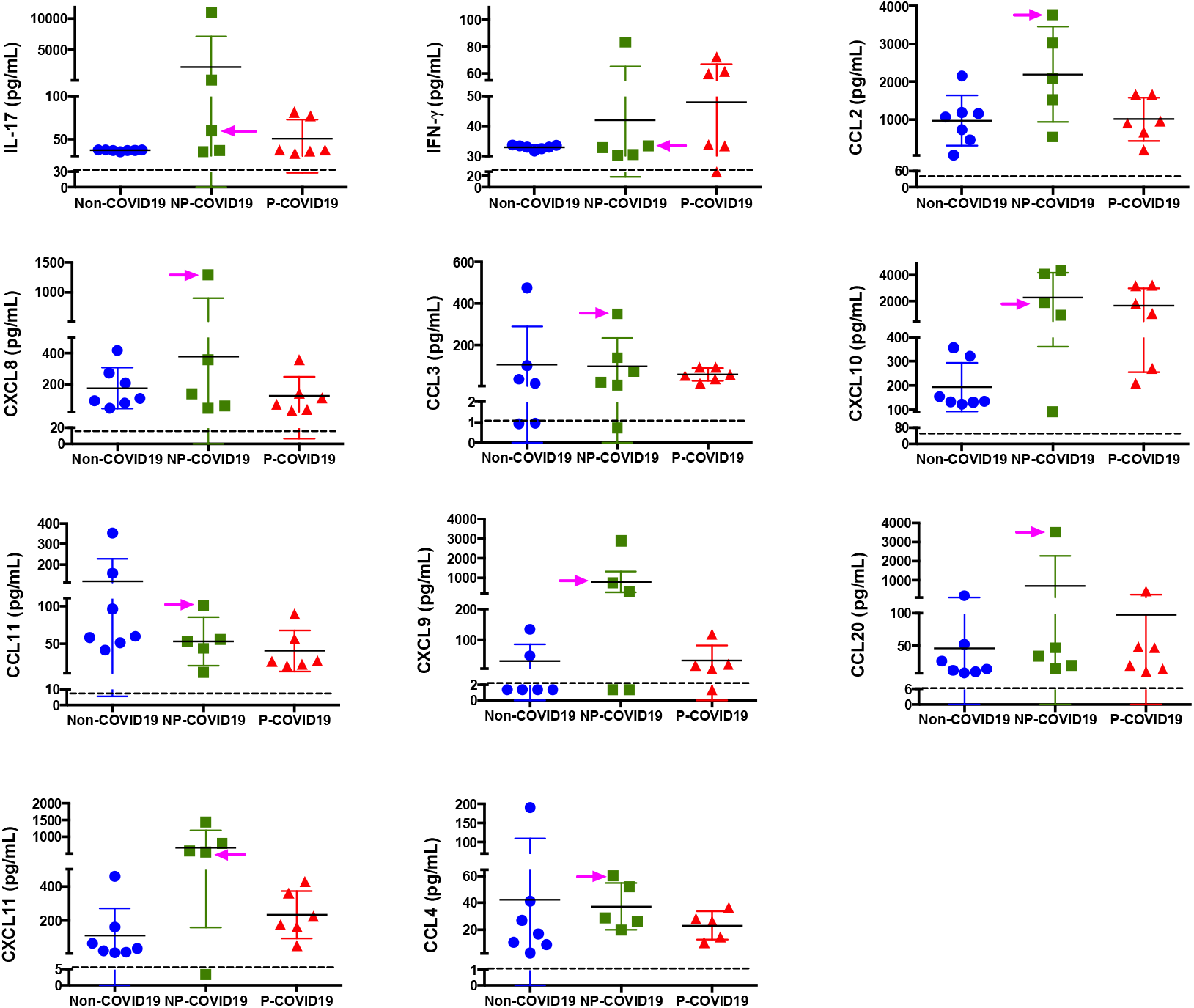
Similar Cytokine and Chemokine serum concentration among groups. Serum was isolated and concentration of cytokines and chemokines was determined using bead-based immunoassays as described in methods. Results are expressed as mean ± SD. Significance value was *p*<0.05. Kruskal-Wallis and Dunn’s multiple comparisons test was calculated. Non-COVID19; Non-pregnant, non-COVID19. (n=7). NP-COVID19; Non-Pregnant-COVID19, (n=5). P-COVID19; Pregnant-COVID19, (n=6). The arrow points the fatal case in NP-COVID19 group.

**Fig 4.**
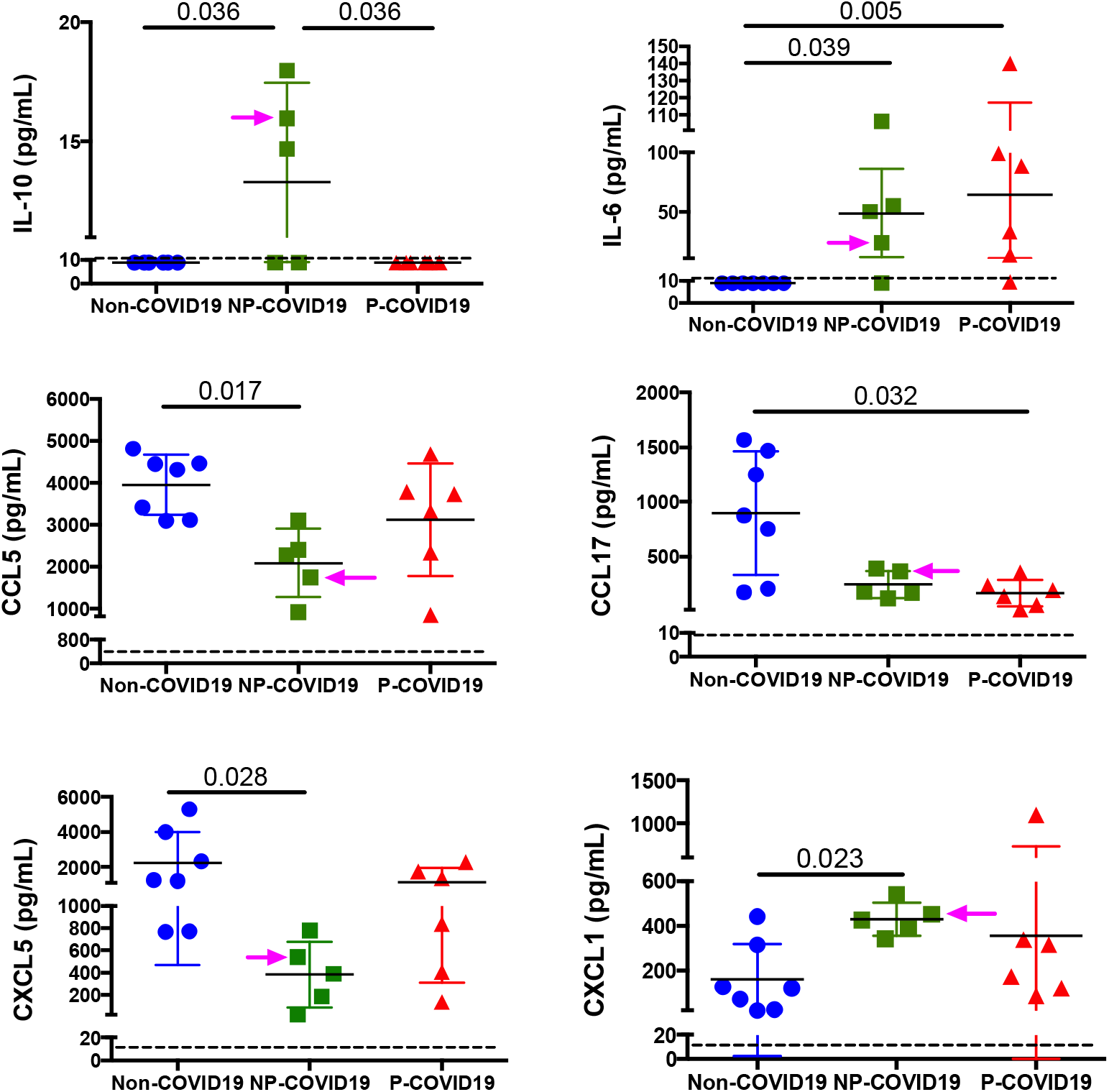
Differential Cytokine and Chemokine serum concentration among groups. Serum was isolated and concentration of cytokines and chemokines was determined using bead-based immunoassays as described in methods. Results are expressed as mean ± SD. The dash line represents the limit for detection. Significance value was considered *p*<0.05. Kruskal-Wallis and Dunn’s multiple comparisons test was calculated. Non-COVID19; Non-pregnant, non-COVID19 (n=7). NP-COVID19; Non-Pregnant-COVID19 (n=5). P-COVID19; Pregnant-COVID19 (n=6). The arrow points the fatal case in NP-COVID19 group.

## Discussion

Six coronaviruses (CoV) are capable of infecting humans and causing respiratory disease, among them the Severe Acute Respiratory Syndrome (SARS-CoV) and the Middle East Respiratory Syndrome (MERS-CoV) coronavirus show high fatality rate in pregnant women (25 and 35% respectively) (27, 33). SARS and MERS also lead to premature delivery, intensive care treatment for newborns, and perinatal death. In contrast, despite the high virulence of COVID19, few cases have been reported in pregnant women around the world, and no maternal deaths have been reported (34-37).

Case reports have been published for pregnant women infected with SARS-CoV-2 (38-40), and also as retrospective studies and systemic reviews that shows a similar clinical characteristics and outcomes like non-pregnant women (34, 37, 41, 42), however, the number of cases seems to be very low around the world. In agreement with these studies we observed that cough, myalgia, nasal congestion among others are similar in frequency by pregnant and non-pregnant women with COVID19, and no significant statistical difference was detected, however clinicians must be alert with minimal signs to early COVID19 diagnosis to avoid complications. In agreement with Lokken et al, we observed a similar frequency of underlying disease for non-pregnant and pregnant women with COVID19 (41). However, obesity is a public health concern in México, and literature mention that this condition is frequently observed in severe or critical cases of COVID19 (3), our study also includes four cases of overweight/obesity. Despite the presence of this underlying condition, no fatal outcome was observed. Future studies must focus on these and other comorbidities to offer early treatment and increase the probability to medical success.

Yan et al reports in the biggest pregnant series that two-third of patients had lymphopenia (37). This condition is also present in non-pregnant patients and is associated with severe and critical cases. Our study showed more cases with abnormal white cell count in the NP-COVID19 group, suggesting that pregnant women can limit the effects of SARS-CoV-2 infection without big change in white blood counts. The Neutrophil/Lymphocyte Ratio (NLR) is an inflammatory biomarker that can be used as an indicator of systemic inflammation in sepsis and COVID19 (43-45), a NLR higher than 5.0 is a risk factor sepsis diagnosis (43) while in patients with COVID19 could be as low as 3.13 or as high as 11.75 (44, 45). Our result show that NP-COVID19 and P-COVID19 groups have a mean value above the cutoff for sepsis, and our only fatal case was above this cutoff. A greater study as series of cases must be analyzed to know if the NLR ratio in pregnant women could be useful to prognostic a better response to COVID19 than general population.

False-negative testing for SARS-CoV-2 RT-PCR is a relevant problem especially in pregnant women, Kelly et al reports that five repeating test until get a positive test (46), these authors mention as sources of variability the anatomic area sampled, quantity of virus, stability of the RNA, timepoint in disease course and assay variability, all of them equally present in pregnant patients, and lead to risk situation to other patients, including healthcare workers (47), similar situation could be also for IgM and IgG antibodies positive test, indicating that concurrent test between RT-PCR and antibodies test could be a better option to an early diagnosis of COVID19 in pregnant women. We detected that serum IgM was in 66% of patients in the group of P-COVID19 while none of the women in the NP-COVID19 group was positive indicating that IgM test could be useful to recognize positive cases of COVID19 in pregnant women, more studies are necessary to confirm or discard this idea.

The major source of cytokine and chemokine in COVID19 non-pregnant patients are the epithelial and endothelial cells plus macrophages and NK or NKT cells, but in pregnant women the placenta is an alternative source for these molecules (48). Early in the pregnancy the maternal-fetal interface is high dependent of communication by the chemokine network and placenta is a chemokine source that it is strictly regulated trough the gestation (48, 49). Chemokines such as CCL2, CCL4, CCL5, CXCL8 and CXCL10 is higher expressed in healthy term labor than in pregnant women without labor (50), this is an important issue to understand because may be the cytokine storm reported in COVID19 patients could alter the physiologic progress in labor and increasing the morbidity in pregnant women.

We observed that certain cytokine and chemokine in COVID19 pregnant women are similar in concentration than in patients in the NP-COVID19 group (Figures 2). Huang et al., reported the plasma concentration for IL-17, IFN-γ, CCL2, CXCL8, CCL3, CXCL10, and CCL11 in patients COVID19 non-pregnant in the ICU care and in the No-ICU care, our results are quite similar to Huang’s report, specially the pattern of cytokine-Chemokine as group and the expression for each cytokine and chemokine individually is reminiscent between reports. Also, we noted that concentration reported by us is between 5 to 10 times higher than in the Huang’s report, probably because we did assess using serum instead plasma, despite this quantitative difference the distribution of cytokine and chemokine are quite similar. This cytokine storm in COVID19 share some characteristics with the Macrophage Activation Syndrome (MAS), this similarity could be taken into considerations for medical therapeutics (like anti-IL-6) in pregnancy (5). In contrast some cytokines and chemokines shown differences between Huang’s report and ours. IL-10 is different between NP-COVID19 *vs*. P-COVID19 while Huang’s report shows a similar response between ICU care *vs*. No-ICU care, we observed a higher concentration of IL-6 than the Huang’s report, and again our P-COVID19 group shows an opposite response than No-ICU care group. Huang’s report analyze a n=28 in the No ICU care group, and in the ICU care group n=13, meanwhile our study analyzes between two and four times less patients, precautions must take about our data.

CCL5 shows a different pattern to Huang’s report, both ICU and No-ICU patients are higher expression than control group, and we observed just the opposite with a clear lower CCL5 level for NP-COVID19. We interpreted that P-COVID19 expressed CCL5 as a not a high-risk characteristic for the SARS-CoV-2 infection, this agrees with a very low number of reported cases among pregnant women around the world, and a small number of fatal cases despite high number of infected patients.

To our knowledge this is the first study that show serum cytokine and chemokine concentration in pregnant women with COVID19. Moroever, the Non-COVID19 group is also of interest since they are female health workers exposed to COVID19, our result show that the exposure to SARS-CoV-2 is not sufficient to elicited a cytokine storm despite high stress they have. However, the limitation is the small number of cases for analysis, due to the time when the pregnant patients requested medical attention, and it was not possible to monitor them for a long time.

There are few studies available about the effect of COVID19 in pregnancy and it has published that pregnant women with COVID are not sufficiently taken into account in clinical research (51), especially in immunology. This is important because contrary to assume that pregnancy in COVID19 could lead to high number of fatal outcomes such as SARS (27), despite data shows just the opposite. So, if pregnancy is a condition were the immune system is capable to handle this viral challenge we must to learn more about tolerance in immune response in pregnancy, and apply that knowledge to successful treatment for the non-pregnant patients.

## Contributors

ACV and CLM conceived and designed the study, and contributed to the analysis of data. ACV write the first version of report, and LAAP, EFO contributed to critical revision of the report. BZR, JCBG and GMLGA contributed to clinical evaluation of patients and supervision for medical treatments and care patients. ACV, LAAP, EFO, GLCR, PMC, MTGR, JLPC, SVRA, RMS, ACC, ESR, DSM, RCLB, AHVC, AEP, RRMO, GFP, contributed to data acquisition, data analysis or data interpretation. All authors reviewed the final version. CLM reviewed and approved the final version.

## Data Availability

The authors confirm that the data supporting the findings of this study are available within the article.

## Acknowledgements

This project was supported by the Mexican National Research Council (CONACYT), Project No. 313494 (awarded to CLM). The authors thank the staff at the Specialties Hospital, National Medical Center “ XXI Century”, and Gynecology & Obstetrics Department in the General Hospital of Mexico “ Dr. Eduardo Liceaga”, and to Dr. Iris Estrada for taking the time to read the manuscript and for her critiques and discussion. In memory of Dr. Leopoldo Flores-Romo. We thanks to Myriam Morales, who edited the manuscript in English.

